# Multiple Introductions of Mpox virus to Ireland during the 2022 International Outbreak

**DOI:** 10.1101/2023.09.18.23295695

**Authors:** Gabriel Gonzalez, Michael Carr, Tomás M. Kelleher, Emer O’Byrne, Weronika Banka, Brian Keogan, Charlene Bennett, Geraldine Franzoni, Patrice Keane, Luke W. Meredith, Nicola Fletcher, Jose Maria Urtasun-Elizari, Jonathan Dean, Brendan Crowley, Derval Igoe, Eve Robinson, Greg Martin, Jeff Connell, Cillian F. De Gascun, Daniel Hare

## Abstract

**Background:** mpox (formerly Monkeypox) virus (MPXV) was considered a rare zoonotic disease prior to May 2022, when a global epidemic of cases in non-endemic countries led to the declaration of a Public Health Emergency of International Concern. Previously, mpox infection was associated with symptoms similar to smallpox, although substantially less severe, including fever, an extensive characteristic rash and swollen lymph nodes.

**Aim:** Elucidating the origin and molecular characteristics of the virus circulating in the Republic of Ireland in the period between May and November 2022.

**Methods:** Whole-genome sequencing of all MPXV cases (80%; n=178/219) analysed against sequences from public databases (n=2695). Bayesian approaches were used to infer the divergence time between sequences from different subclades and transmission events involving different countries.

**Results:** The circulating virus belonged to the clade IIb B.1 lineage and, notably, the presence of twelve separable and highly-supported subclades consistent with multiple introductions into the country. Such a hypothesis of multiple importation events was supported further by the estimation of the time to the divergence of subclades. Additionally, inferred MPXV transmissions involving different countries and continents were indicative of an extended international spread. The analysis of the mutations in the Irish sequences revealed 93% of the mutations were from cytosine to thymine (or from guanine to adenine), leading to a high number of non-synonymous mutations across the subclades.

**Conclusion:** In the context of extremely high national sequencing coverage, we provide new insights into the international origin and transmission dynamics supporting multiple introductions into the Republic of Ireland.

**Conflict of Interests:** None to declare.

## Introduction

Mpox (Monkeypox) virus (MPXV) is a double-stranded DNA virus in the genus *Orthopoxvirus* within the *Poxviridae* family (Xiang and White, 2022) and is regarded as a High consequence Infectious Agent (HCID). Despite its name, the natural reservoir of the virus remains unknown but it is transmitted among small mammals, such as rodents (W.H.O., 2023). The length of the MPXV genome is approximately 197.2 kb encoding a predicted 191 proteins, and is genetically closely related to the variola (smallpox) virus. Mpox symptoms can last between two and four weeks and include fever, sore throat, headache, muscle aches, low energy, swollen lymph nodes and rash; however, the combination and severity of these symptoms varies among patients and some severe cases can develop skin lesions (Saxena et al., 2023). The virus is transmitted to humans by direct contact with infected animals, through scratches and bites, or materials, such as contaminated clothes or needles, or by contact with lesions of infected people. Sexual transmission appears to be a major route of transmission, in particular during the recent outbreak, (W.H.O., 2023; Sun et al., 2023).

Two major clades of MPXV have been recognised as circulating endemically in Africa, one in Central Africa (clade I) and the other one in West Africa (clade II) with differing transmissibility and case fatality rates (Quarleri et al., 2022). In this context, an increased number of cases started appearing internationally in May 2022, and were characterised as part of the West African clade II. Based on viral phylogenetic properties and epidemiological evidence of increased human-to-human transmission outside of endemic geographic locations, MPXV cases in the context of the 2022 outbreak were classified within a distinct subclade, termed IIb (Gigante et al., 2022). The outbreak has been reported across multiple countries on different continents and has differed from earlier outbreaks of clade I and IIa, in terms of host age (54.3% of individuals in their thirties), sex (predominantly male), and sexual transmission the most commonly reported (Bragazzi et al., 2023).

The overall case fatality rate of mpox has been estimated at 8.7%, though substantial differences have been observed between clades: estimated as 10.6% for clade I, versus 1-3.6% for clade IIa (Bunge et al., 2022; Americo et al., 2023), and a mortality rate in the range 0.01-1.81% for Clade IIb associated with the 2022 event (Reda et al., 2022). Whilst the 2022 international outbreaks of MPXV clade IIb had milder clinical severity, therefore, it represented a significant public health concern due to the significant increase in reported sexual transmission and concerns regarding “spill over” into other more susceptible populations (Sun et al., 2023; Borges et al., 2023). Due to the unprecedented nature of the mpox epidemic outbreaks outside endemic African countries and its classification as an HCID, the WHO declared a Public Health Emergency of International Concern (PHEIC) in July 2022 (Wenham and Eccleston-Turner, 2022).

MPXV clade case designation by genomic analyses facilitates an evidence-based assessment of transmissibility and the ability to distinguish new importation events. To establish a comprehensive genomic epidemiological baseline for national surveillance in the Republic of Ireland, whole-genome sequencing (WGS) was performed at the National Virus Reference Laboratory for all available MPXV laboratory-confirmed positive cases. Genome sequences (n=178 sequences) were obtained covering an extremely high percentage (>80% of cases) of all cases between May and November 2022 (n=219 cases). This comprehensive MPXV genomic dataset was analysed further to elucidate the dynamics of the 2022 MPXV epidemic in the Republic of Ireland to ascertain whether single or multiple introductions of the virus were responsible.

## Materials & Methods

### Sample collection

Samples (predominantly swabs of skin lesions/pustules transported in virus transport media, VTM)) were received from 219 symptomatic individuals who presented for medical attention. Following lysis and inactivation in a Containment Level 3 (CL3) facility with a VTM to guanidinium isothiocyanate lysis buffer mixed in a ratio of 1:2.5, total nucleic acids were extracted on the Roche MagNA Pure 96 platform as follows: 200 µL of swab material in VTM was added to 500 µL Roche lysis buffer, and, following external lysis, were extracted on the Roche MagNA Pure 96 system (input volume 450 µL and eluted in 100 µL). Alternatively, DNA from 200 µL of material in VTM was extracted using the Qiagen QIAamp DNA Mini kit, according to the manufacturer’s instructions.

### Molecular Diagnostics

Real-time PCR was performed on the ABI7500 SDS platform employing initially the pan-*Orthopoxvirus* zoonotic (non-variola) RT-PCR kit (Altona); for logistical reasons the assay was switched to the MPXV generic assay described by Li et al. (Li et al., 2010). However, a CDC notification of a rare but significant deletion in the target TNF gene motivated to change the assay to the LDT pan-*Orthopoxvirus* assay currently in use (Schroeder and Nitsche, 2010).

### Whole-Genome Sequencing

MPXV qPCR positive samples were processed for WGS following a tiled amplicon approach and next-generation sequencing (NGS) approach employing Oxford Nanopore Technologies (ONT). Tiled amplicons were generated from each sample employing two separate MPXV primer pools (P/N 50025, MBS), according to the manufacturer’s instructions (Matthijs Welkers et al., 2022). Amplicons were barcoded employing the Rapid Barcoding Kit 96 (SQK-RBK110.96; ONT) and loaded onto a R9.4.1 flow cell (FLO-MIN106D; ONT) with 23 samples and a negative control on each run. The data generated were basecalled and demultiplexed with Guppy v5.1.13 (ONT). The sequence reads were trimmed to remove adapters and barcoded sequences with Porechop v.0.3.2pre (https://github.com/rrwick/Porechop). NGS reads were assembled employing a MPXV genome sampled in November of 2021 in the United States of America (GenBank ON676708) as the reference. The assembled consensus genome sequences were deposited in GenBank (https://www.ncbi.nlm.nih.gov/genbank/) under accession numbers OR146268-OR146445 and also in GISAID (Supplementary Table S2). MPXV clades were assigned following the automatic assignment by GISAID after data submission.

All multiple sequence alignments (MSA) were performed with MAFFT by the algorithm FFT-NS-1 (Katoh et al., 2019). Phylogenies were inferred using a maximum-likelihood (ML) approach with RAxML using the general time reversible (GTR) substitution model and support for the branches was estimated with a bootstrap approach considering 500 repetitions (Stamatakis, 2014). MSA included ON676708 as reference to annotate the coding regions and to analyse the effects of the nucleotide mutations as amino acid substitutions in the encoded proteins with only mutations on nucleotides supported by sequence depth > 20 reads analysed further.

### Inferring time to the most recent common ancestor

The divergence times of the different MPXV subclades to the most recent common ancestor (tMRCA) were inferred using BEAST2 v2.5.7 (Bouckaert et al., 2019). The reference ON676708 was employed as an outgroup for rooting the tree. The substitution model was chosen using the model test in RAxML GUI (Edler et al., 2021) and the Bayesian information criterion (BIC), choosing the Hasegawa-Kishino-Yano (HKY) substitution model as the best fit for the sequences requiring the least amount of parameters with a proportion of invariant sites (+I). The different models with a constant clock or an uncorrelated Optimised Relaxed Clock (ORC) (Douglas et al., 2021), with the population modelled either as constant size, exponential growth or Bayesian skyline, were considered and compared to identify the most likely model among the six combinations. All models were calibrated with the collection date of the samples. Each model was executed for 10^8^ steps sampling every 10^4^ to assure an Effective Sample Size (ESS) > 100 for all parameters in the models, i.e. the number of effectively independent draws for each parameter from the posterior distribution that the Markov chain was equivalent to. The model parameters were summarised and explored with Tracer 1.7.1, and tree of the best model was summarised using the ‘treannotator’ programme in the BEAST bundle to extract the divergence time and the mutation rate of the subclades.

### Testing association with international MPXV genome sequences

Contemporaneous sequences were downloaded from the MPXV database in GISAID (https://gisaid.org/) with the filtering criteria requiring near complete genome sequences and high quality (>90% genome coverage) (Supplementary Table S1). Additionally, reference sequences from previous MPXV outbreaks were downloaded from GenBank (Supplementary Table S1). The association of Irish genome sequences to sequenced cases from other countries was tested by separating the group of Irish sequences into the following twelve IIb subclades: B.1, B.1.1, B.1.2, B.1.3, B.1.5, B.1.7, B.1.8, B.1.9, B.1.10, B.1.11, B.1.12, and B.1.15. The MSAs of these groups included GISAID sequences from the same subclades to test the phylogenetic relationships and putative origins of the MPXV with the Bayesian inference approach of Outbreaker2 (Campbell et al., 2018), using the collection date reported in GISAID and assuming an infectious period of 28 days with the highest probability of transmission between days 6 and 15, following the description of cases and transmission via skin lesions (W.H.O., 2023). The assumed mutation rate was 5×10^−5^, the lower end of the mutation rate range from the BEAST analysis. The Outbreaker2 inference was run for 10^6^ steps in each of the eleven groups, with 10% burn-in and sampling every 2500 steps. Among the Outbreaker2 results of transmissions, we considered as evidence of transmission between two samples those links with support > 0.3 and suggesting less than two generations.

### Ethical approval

The analysis of the data was approved by the University College Dublin Human Research Ethics Committee – Sciences analysed data with the Research Ethics Reference Number: LS-LRSD-23-197-Gonzalez-Hare.

## Results

### Characteristics of the 2022 MPXV outbreak

The 219 laboratory-confirmed MPXV qPCR-positive samples analysed in the present study were collected between May and November 2022. As reported by the Health Service Executive (HSE) of the Republic of Ireland (ROI), the majority of cases were males with a median age of 35 years with 79.8% aged between 18-44 years (HPSC, 2023). These cases were distributed over time with the peak in August (> 60 cases) (Fig. 1).

**Figure 1.**
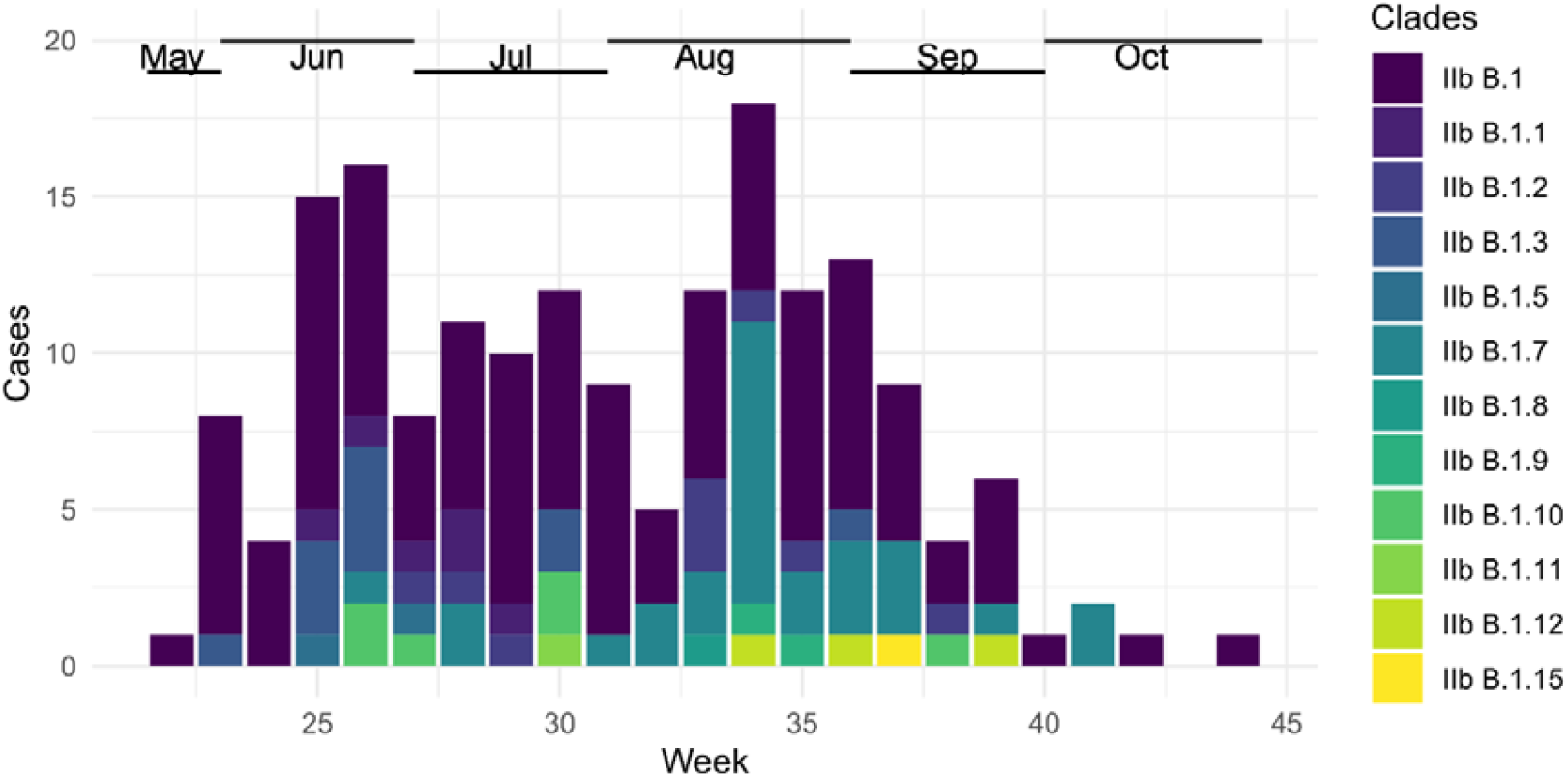
MPXV cases reported in the Republic of Ireland. The horizontal and vertical axes represent the epidemiological week of 2022 when the samples were received and the total number of reported cases across the country, respectively. The distribution of subclades assigned by GISAID has been coloured according to the legend. Months are shown on the top of the panel.

### WGS of Irish MPXV in 2022

Among the 219 cases, 178 (80%) were successfully sequenced, of which 175 met the ≥90% coverage threshold with an average genome coverage of 99.3 ± 2% (Supplementary Table S2). The high coverage of the sequences facilitated the mutational analysis compared to the MPXV clade IIb reference genome ON676708, which was chosen as reference. The transition/transversion ratio was 22.8, with 286 positions showing mutations relative to the reference genome. Of the total number of mutations, 47% and 46% were transitions C to T and G to A, respectively (Supplementary Table S3). Relative to the reference genome annotation, these mutations were approximately seven times more frequent in coding regions than in the non-coding regions (Supplementary Table S4).

Moreover, 154 mutations were non-synonymous mutations while 100 were synonymous mutations with 32 characterised as non-coding mutations (Supplementary Table S4). These mutations distributed into the different GISAID-assigned clades as follows: a total of 11 mutations were considered common to all sequenced samples with their presence in >90% of the sequenced samples; 194 mutations were identified as isolated single nucleotide variants (SNV) present in only one sequence, among them, 132 (n=105) were in the IIb B.1 subclade (dark green sequences in Fig. 2). Conversely, a total of 104 SNVs were distributed and characteristic to each subclade (Fig. 2), i.e. common to all the sequences under the same subclade in the format IIb B.1.* as follows - B.1.1: 6 SNV (n=6), B.1.2: 20 (n=9), B.1.3: 10 n=11), B.1.5: 7 (n=2), B.1.7: 27 (n=28), B.1.8: 8 (n=1), B.1.9: 5 (n=2), B.1.10: 13 (n=6), B.1.11: 4 (n=1), B.1.12: 2 (n=3), and B.1.15: 2 (n=1). The remaining 28 mutations corresponded to variants within each subclade. These SNV have accumulated following divergence among subclades and point mutations were still accumulating, as was most evident in the basal subclade IIb B.1.

**Figure 2.**
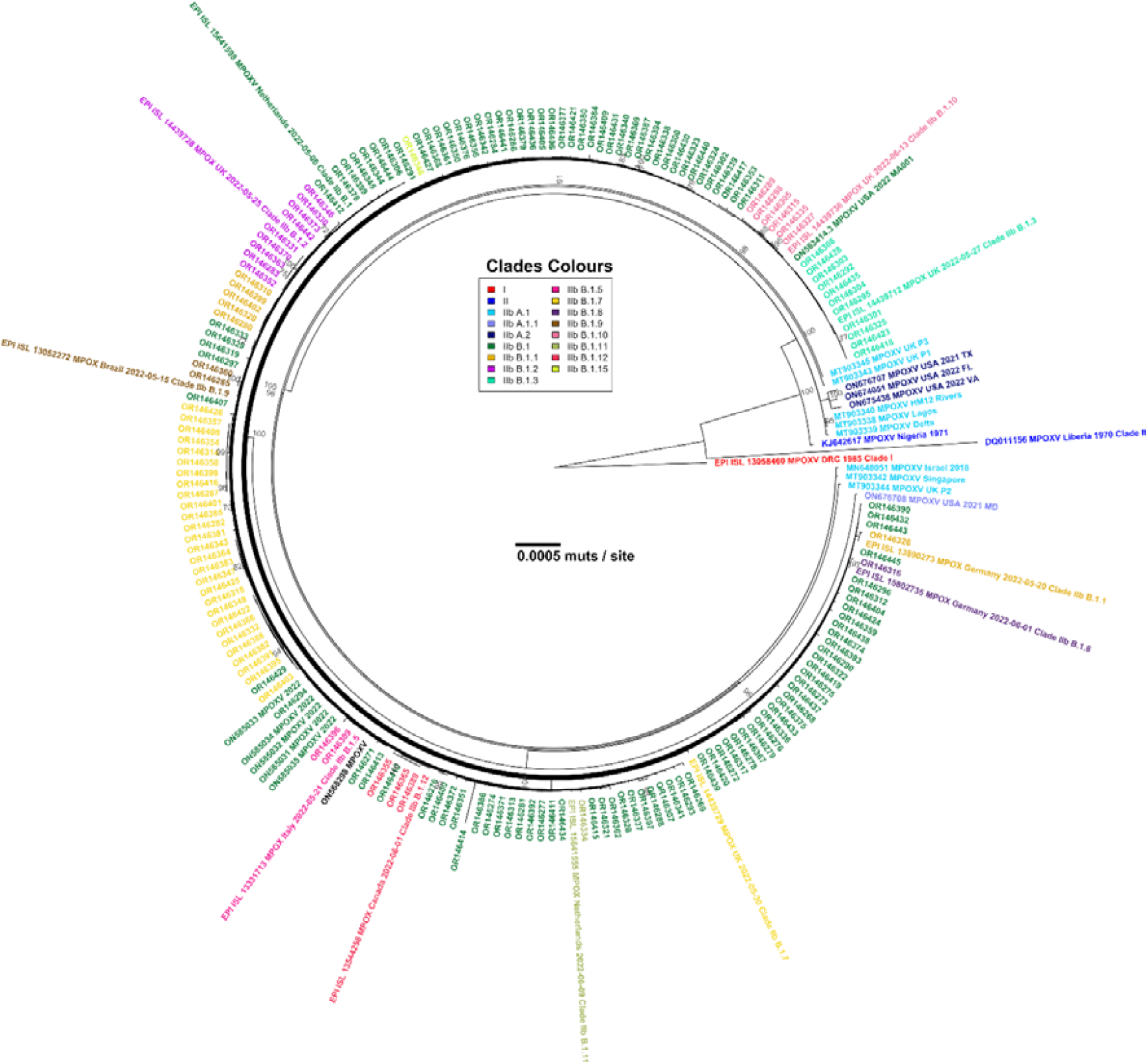
The MPXV phylogenetic tree for whole-genome sequences of Irish cases and reference genomes. The tree was rooted with an MPXV Clade I sequence (GISAID acc. number: EPI_ISL_13058460) and other reference sequences were added to corroborate the clade assignments for the sequences. All external sequences show the corresponding accession number and short description of their origin. Support for branching nodes, estimated with a bootstrap approach (500 repetitions), is shown for support ≥70%. Tip names have been coloured according to the subclade as shown on the colour legend. The scale of the tree represents 5×10^−4^ mutations per nucleotide site.

The subclades assigned by GISAID, the number of characteristic mutations per subclade, and the phylogenetic relationships among our samples and sequences from previous outbreaks confirmed the identity of the cases as belonging to the MPXV clade IIb B.1 (Fig. 2). Furthermore, at least twelve phylogenetically separable and highly-supported subclades were suggested to circulate simultaneously in ROI during the 2022 epidemic (Fig. 2). The cases identified during the outbreak belonged to the subclade IIb B.1, however, during the 20 weeks period when these cases were detected, distinct variants were identified suggesting at least two alternative hypotheses to explain these findings: (i) the twelve subclades were introduced to ROI in a single initial wave that evolved locally, or (ii) a flow of new subclades to the country in multiple separate importation events.

### Dating the divergence of the Irish MPXV subclades

To investigate further the possible origins of the multiple subclades circulating in ROI, the divergence times of the different subclades were inferred employing a Bayesian approach. The comparison of the considered Bayesian models demonstrated that a combination of a Bayesian skyline modelling of the population growth with an uncorrelated relaxed clock was the model with the highest likelihood for the observed genomic data (Table 1). The dating of the tMRCA of the different subclades to mid-May (2022.37, 95% HPD [2022.28, 2022.40], corresponding to epidemiological weeks 15 and 21 of 2022, respectively) was consistent with divergence prior to the outbreaks in the ROI (Table 1, Fig. 3). This also provided evidence in support of the hypothesis that multiple introductions, not a single importation event, of new and diverse variants into Ireland occurred throughout the 2022 outbreak event. Furthermore, while other phylogenetic groups were observed to have diverged as early as the last week of May, at least two clades, namely IIb B.1.9 and B.1.12, diverged in August 2022 and, given that no earlier cases assigned to these clades were detected, it is considered more likely they were introduced into the country later in the course of the outbreak. Additionally, other sub-clusters in IIb B.1 and B.1.2 are suggested to have diverged after August and as late as September 2022 (Fig. 3). It is noteworthy that despite the best fit being a Bayesian skyline with the ORC, all the inferred models had very similar dates for the tMRCA and relatively high mutation rates in the range [7.2E-5, 1.3E-4] which is notably higher than the 10^−6^ mutation rate previously reported for the family *Orthopoxviruses* (Babkin et al., 2022).

**Table 1.**
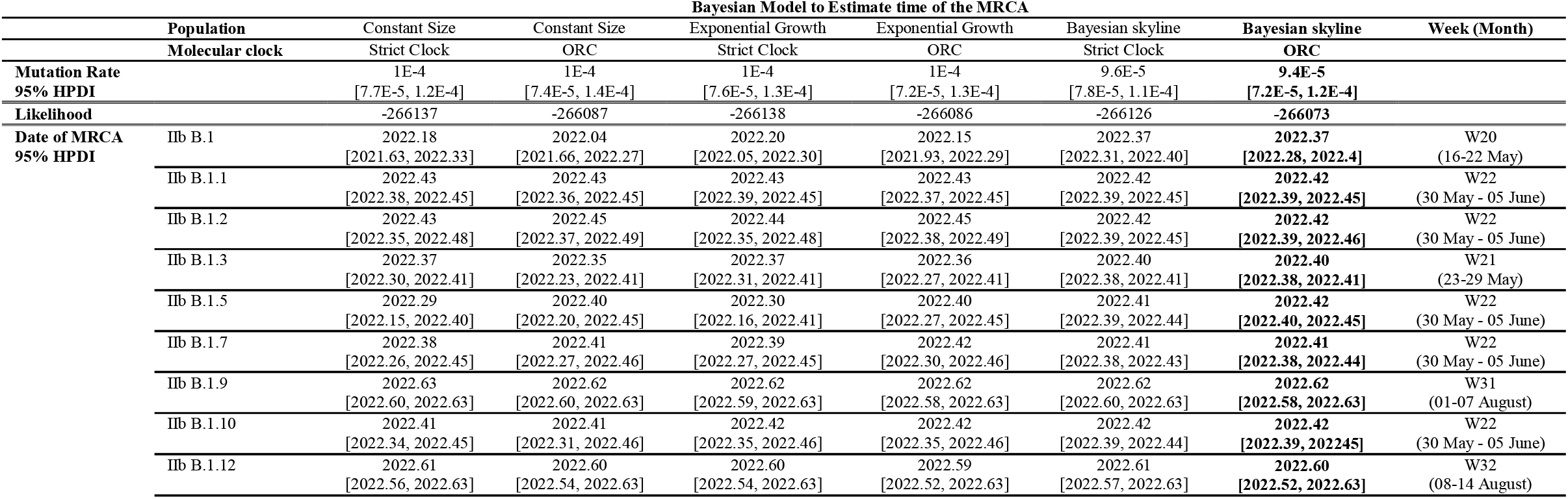
Comparison of Bayesian models and the inferred time to the MRCA of the MPXV clades in the Republic of Ireland.

|  |  | Bayesian Model to Estimate time of the MRCA |  |  |  |  |  |  |
| --- | --- | --- | --- | --- | --- | --- | --- | --- |
| Population |  | Constant Size | Constant Size | Exponential Growth | Exponential Growth | Bayesian skyline | Bayesian skyline | Week (Month) |
| Molecular clock |  | Strict Clock | ORC | Strict Clock | ORC | Strict Clock | ORC |  |
| Mutation Rate |  | 1E-4 | 1E-4 | 1E-4 | 1E-4 | 9.6E-5 | 9.4E-5 |  |
| 95% HPDI |  | [7.7E-5, 1.2E-4] | [7.4E-5, 1.4E-4] | [7.6E-5, 1.3E-4] | [7.2E-5, 1.3E-4] | [7.8E-5, 1.1E-4] | [7.2E-5, 1.2E-4] |  |
| Likelihood |  | -266137 | -266087 | -266138 | -266086 | -266126 | -266073 |  |
| Date of MRCA<br>95% HPDI | IIb B.1 | 2022.18 | 2022.04 | 2022.20 | 2022.15 | 2022.37 | 2022.37 | W20 |
|  |  | [2021.63, 2022.33] | [2021.66, 2022.27] | [2022.05, 2022.30] | [2021.93, 2022.29] | [2022.31, 2022.40] | [2022.28, 2022.4] | (16-22 May) |
|  | IIb B.1.1 | 2022.43 | 2022.43 | 2022.43 | 2022.43 | 2022.42 | 2022.42 | W22 |
|  |  | [2022.38, 2022.45] | [2022.36, 2022.45] | [2022.39, 2022.45] | [2022.37, 2022.45] | [2022.39, 2022.45] | [2022.39, 2022.45] | (30 May - 05 June) |
|  | IIb B.1.2 | 2022.43 | 2022.45 | 2022.44 | 2022.45 | 2022.42 | 2022.42 | W22 |
|  |  | [2022.35, 2022.48] | [2022.37, 2022.49] | [2022.35, 2022.48] | [2022.38, 2022.49] | [2022.39, 2022.45] | [2022.39, 2022.46] | (30 May - 05 June) |
|  | IIb B.1.3 | 2022.37 | 2022.35 | 2022.37 | 2022.36 | 2022.40 | 2022.40 | W21 |
|  |  | [2022.30, 2022.41] | [2022.23, 2022.41] | [2022.31, 2022.41] | [2022.27, 2022.41] | [2022.38, 2022.41] | [2022.38, 2022.41] | (23-29 May) |
|  | IIb B.1.5 | 2022.29 | 2022.40 | 2022.30 | 2022.40 | 2022.41 | 2022.42 | W22 |
|  |  | [2022.15, 2022.40] | [2022.20, 2022.45] | [2022.16, 2022.41] | [2022.27, 2022.45] | [2022.39, 2022.44] | [2022.40, 2022.45] | (30 May - 05 June) |
|  | IIb B.1.7 | 2022.38 | 2022.41 | 2022.39 | 2022.42 | 2022.41 | 2022.41 | W22 |
|  |  | [2022.26, 2022.45] | [2022.27, 2022.46] | [2022.27, 2022.45] | [2022.30, 2022.46] | [2022.38, 2022.43] | [2022.38, 2022.44] | (30 May - 05 June) |
|  | IIb B.1.9 | 2022.63 | 2022.62 | 2022.62 | 2022.62 | 2022.62 | 2022.62 | W31 |
|  |  | [2022.60, 2022.63] | [2022.60, 2022.63] | [2022.59, 2022.63] | [2022.58, 2022.63] | [2022.60, 2022.63] | [2022.58, 2022.63] | (01-07 August) |
|  | IIb B.1.10 | 2022.41 | 2022.41 | 2022.42 | 2022.42 | 2022.42 | 2022.42 | W22 |
|  |  | [2022.34, 2022.45] | [2022.31, 2022.46] | [2022.35, 2022.46] | [2022.35, 2022.46] | [2022.39, 2022.44] | [2022.39, 2022.45] | (30 May - 05 June) |
|  | IIb B.1.12 | 2022.61 | 2022.60 | 2022.60 | 2022.59 | 2022.61 | 2022.60 | W32 |
|  |  | [2022.56, 2022.63] | [2022.54, 2022.63] | [2022.54, 2022.63] | [2022.52, 2022.63] | [2022.57, 2022.63] | [2022.52, 2022.63] | (08-14 August) |

**Figure 3.**
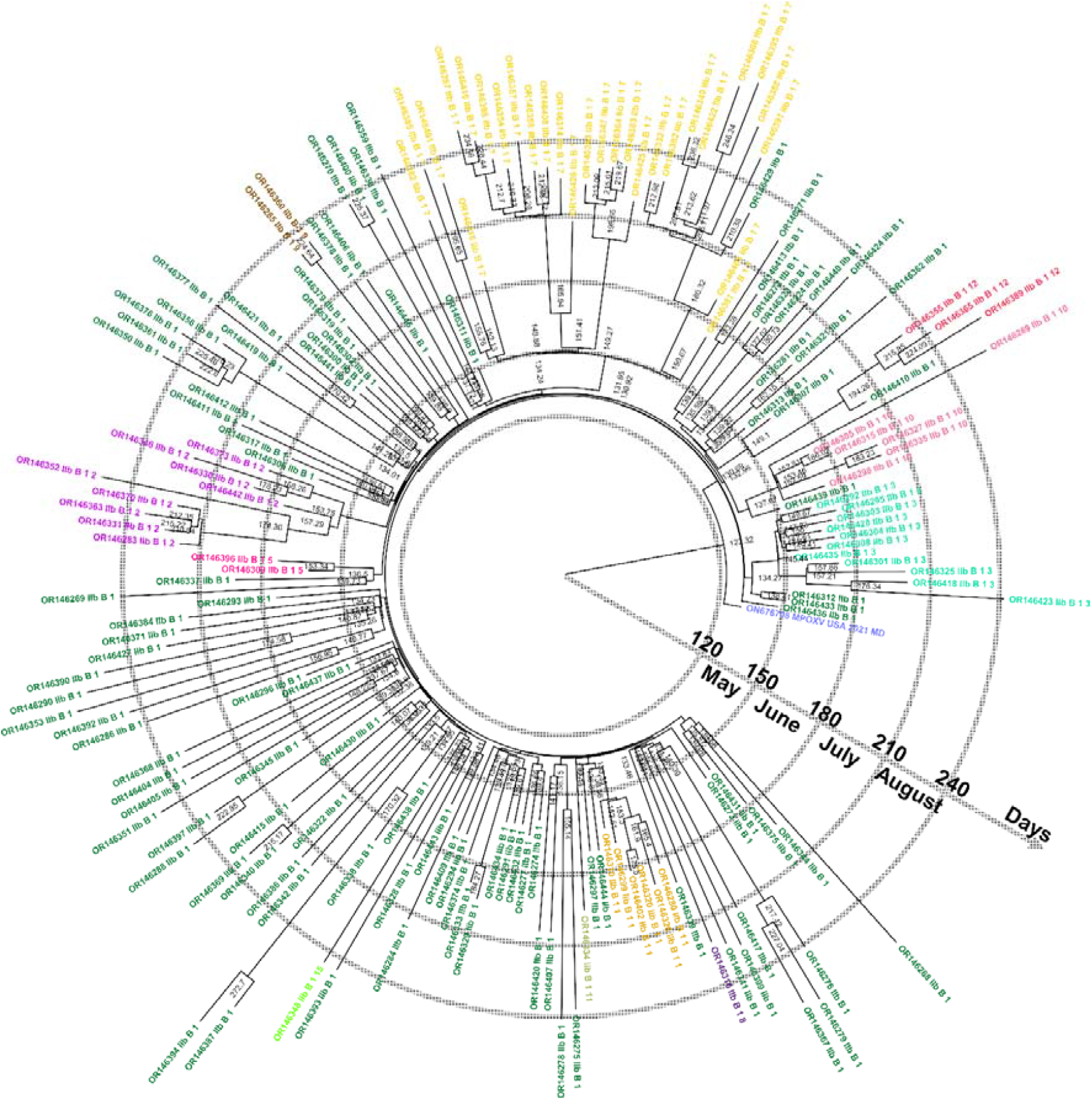
Bayesian inferred phylogenetic tree with the most likely time of the MRCA for the different clades in the Republic of Ireland. The circular tree represents the 175 ROI sequences rooted with ON676708 (IIb A.1). The nodes show the most likely day of the year 2022 for the MRCA, the scale for these days and the corresponding months are shown as blue concentric circles. Clades are coloured as in Fig. 2 and have been added to the tip names for reference.

### Uncovering possible origins of the MPXV introductions to Ireland

As mentioned above, the clade IIb subclade B.1 was predominant during the 2022 MPXV outbreak in ROI (n=105), and consistent with the distribution in other locations for the same time period with between 33% and 74% of sequences from this subclade (Table 2). As some of the other circulating subclades were suggested to have diverged some time prior to the first MPXV case detected in the ROI (31 May 2022), the subclade sequences were compared with sequences from other geographical locations to identify putative origins for the different clades and to explore further the evidence for any transmission events within these individuals (Fig. 4, Supplementary Fig. S1 and Supplementary Fig. S2). Despite the conservative approach used to infer the transmission, requiring less than two generations and a support of > 30% in the Outbreaker2 inference, many nodes were connected across the different countries and continents, reflecting the international transmission dynamics. Many of the inferred infectious links supported the transmission between cases from the same geographical location as community transmission in IIb B.1.9 and B.1.10 (Supplementary Fig. S2C-D) or the more numerous cases in IIb B.1.1, B.1.2 and B.1.7 (Supplementary Fig. S1A, B, D). The Bayesian analysis inferred strong support for importation events based on the collection dates and phylogenetic relationships among the sequences. The inference suggested importations into ROI from both Europe (United Kingdom, Portugal, Germany, Switzerland) and North and South America (the United States of America, Colombia, and Brazil) (Fig. 4).

**Table 2.** MPXV clade IIb subclades in GISAID from the 2022 outbreak.

| Subclades | Location |  |  |  |  |  | Cases |
| --- | --- | --- | --- | --- | --- | --- | --- |
|  | Ireland | Europe | North America | South America | Asia | Oceania |  |
| **IIb B.1** | 60% | 74% | 60% | 61% | 33% | 100% | **1874** |
| **IIb B.1.1** | 3% | **7%** | 4% | 22% | 11% |  | **217** |
| **IIb B.1.2** | 5% | 3% | 12% | 2% | 22% |  | **195** |
| **IIb B.1.3** | 6% | 3% | 4% |  | **26%** |  | **102** |
| **IIb B.1.5** | 1% | 3% | 1% |  | 4% |  | **56** |
| **IIb B.1.7** | 16% | 4% | 5% | 1% | 4% |  | **137** |
| **IIb B.1.8** | 1% | 2% | 3% |  |  |  | **51** |
| **IIb B.1.9** | 1% | 1% | 0% | 4% |  |  | **31** |
| **IIb B.1.10** | 3% | 1% | 0% | 8% |  |  | **50** |
| **IIb B.1.11** | 1% | 0% | 5% | 1% |  |  | **65** |
| **IIb B.1.12** | 2% | 1% | 4% | 1% |  |  | **50** |
| **IIb B.1.15** | 1% | 1% | 0% | 1% |  |  | **14** |
| **Cases** | **175** | **1253** | **1074** | **312** | **27** | **1** |  |

**Figure 4.**
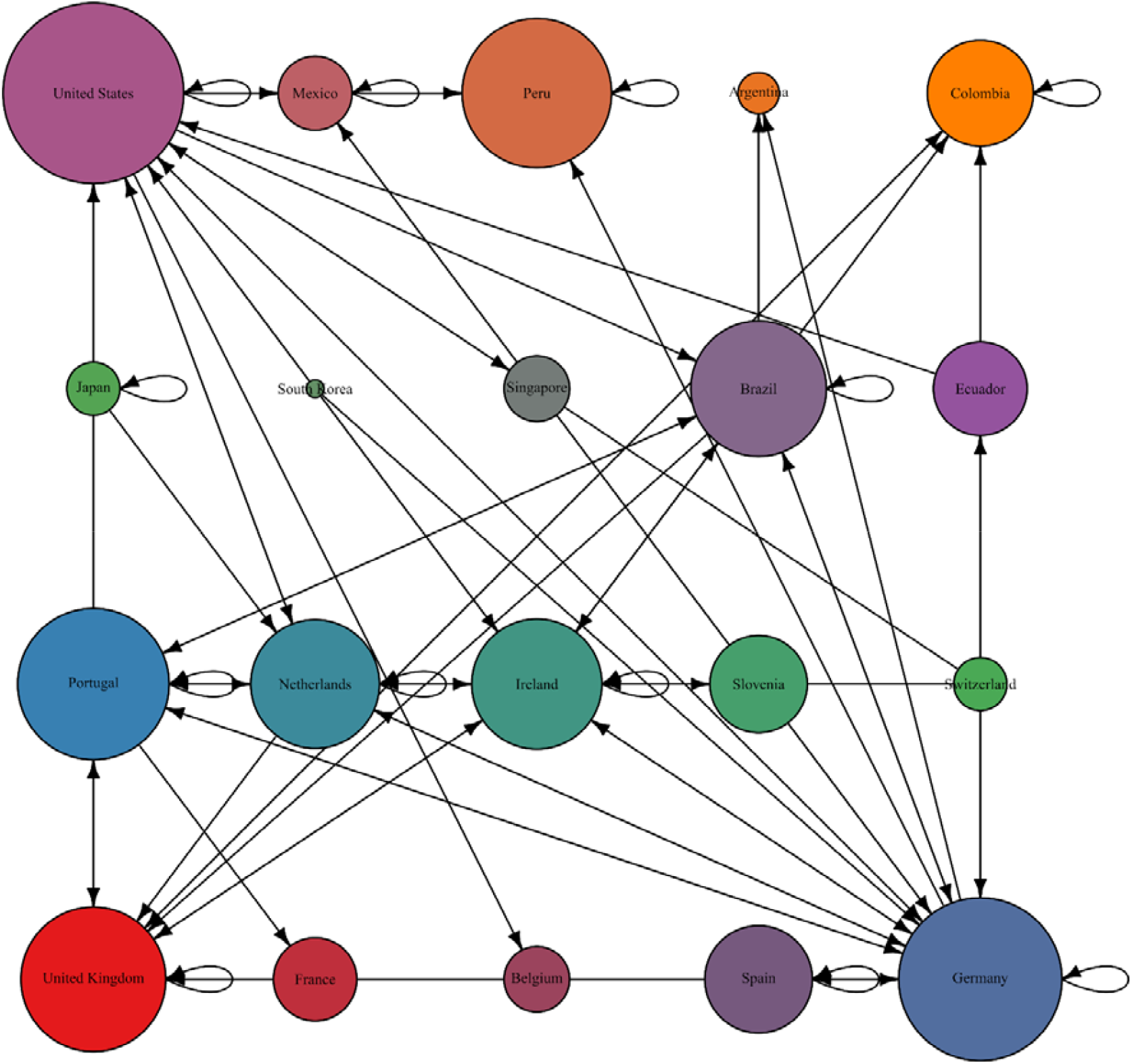
Summary of inferred transmissions among countries detected in the subclades detected in the Republic of Ireland. The nodes represent countries where the lineages of subclades in the clade IIb B.1 were sequenced. The size of the nodes reflects the number of sequences reported by each country. The arrows represent the direction of inferred transmission events.

## Discussion

The first detections of MPXV in Ireland was notified on May 31^st^ 2022 with the earliest onset of symptoms on May 13^th^. The major peak of cases was in and August 2022 and declined significantly since this time. A similar observation has been shared in other jurisdictions where the numbers of cases have decreased significantly worldwide. The explanation for the decline in case numbers is likely multifactorial including effective public health communication regarding protective measures, the subsequent introduction of safe practices to mitigate against transmission in vulnerable groups. Parallel with this the introduction of targeted vaccination policy to protect those at most risk and the reduced mass gathering events in late 2022 (HSE, 2022; Yang et al., 2023). The smallpox vaccine was offered as MPXV prophylaxis and it has been suggested that the previous cessation of the smallpox vaccination around the world could have been one of the factors contributing to the emergence of the current outbreak (Bunge et al., 2022).

Since the 2022 outbreak in the ROI described here, there have been further sporadic cases (n=5) notified in December 2022, January 2023 and August 2023, and internationally a surge of cases in East Asia during July and August (Murphy, 2023). This indicates importation events are likely occurring with the potential for onward transmission. Such cases highlight the need for vigilance, early detection with timely genomic surveillance and rapid public sharing of sequences to monitor for potential amino acid substitutions and for MPXV clade changes over time on both national and international levels. The availability of the large genomic epidemiological baseline from 2022 described in this study will facilitate the delineation of transmission patterns and, specifically, the ability to distinguish imported from autochthonous transmission events as the priority must be to mitigate against further disease transmission by preventing the establishment of recalcitrant reservoirs arising from potential silent transmission, i.e. in asymptomatic cases (Mizushima et al., 2023), in different geographical regions which will seed further outbreaks.

Our results of MPXV genomic epidemiological analysis demonstrates multiple introductions of the virus to the ROI with evidence for continued divergence during 2022. Such results were consistent with the national published reports documenting that 50.9% (n=116/190) of cases were associated with a recent history of international travel in the 21 days prior to symptom onset (HPSC, 2023). Our analyses also contextualised the Irish cases into the global genomic epidemiology of the unprecedented international MPXV 2022 outbreak (Isidro et al., 2022), with the presence of subclades with higher prevalence in different continents suggested to be transmitted to cases in Ireland.

The dominant clade of circulating MPXV in Ireland was suggested to accumulate mutations at a rate that was at least an order of magnitude higher than previously reported (Babkin et al., 2022), which is in keeping with recent data from a number of other studies (Yadav et al., 2022; Luna et al., 2022). The Bayesian-inferred population model suggested an increase in the effective population size early on in the outbreak that gave origin to many of the different subclades, as a probable consequence of the extensive human-to-human transmission events and adaptation and modification by the human host immune response (Yang et al., 2023). Such results could explain the distinctive pattern of the increases in the number of accumulated mutations as the virus was replicating and being transmitted. The analysis of mutational patterns amongst our MPXV cases strongly supported the hypothesis that much of this divergence is driven by the effects of enzymes of the APOBEC cytosine deaminase family where C to T (or G to A on the lagging strand) mutations predominated (Isidro et al., 2022), as was also observed in the evolution of SARS-CoV-2 during the COVID-19 pandemic (Mourier et al., 2021). Therefore, while MPXV may still be considered a zoonotic infection (W.H.O., 2023), viral phylogenetic and Bayesian analysis of local and international transmission dynamics provide evidence in support of sustained adaptation of the virus to the human and a potential risk to public health by altered transmission (Yang et al., 2023; Luna et al., 2022)

The extremely high sequencing coverage in our study facilitated robust phylogenetic inferences to be drawn by reducing uncertainty as to the direction and dating of the transmission between cases. With lower national genomic surveillance, uncertainty increases and the number of possible transmission routes and epidemiological variables also grows exponentially. As a consequence, this limits the amount of information that can be drawn from the gathered data. The combination of epidemiological data with high sequencing coverage for a particular outbreak can lead to insights into the origins, the transmission dynamics and even susceptibility of different populations as was implemented during the COVID-19 pandemic in tracing the epidemiological dynamics of SARS-CoV-2 (Lucey et al., 2021; Hare et al., 2021).

In conclusion, through detailed phylogenetic analysis of MPXV cases, in the context of extremely high sequencing coverage by international comparison performed in the Irish population, we provide evidence for multiple introductions during the 2022 outbreak underscoring the possibility that importation events could recur here and elsewhere posing a continued threat to public health. This dataset provides an epidemiological baseline to monitor future MPXV importation events by analogy to that employed for other pathogen genomics efforts including influenza, measles and polio. The surveillance of MPXV genetic diversity also enables the assessment of the effectiveness of interventions to contain epidemic threats and the rational development of target-specific prophylactic vaccines. Furthermore, with a better understanding of the nature of MPXV introductions into Ireland, and their subsequent transmission dynamics, evidence based public health interventions can be planned and implemented, and mathematical modelling of mpox epidemiology can inform vaccination policy.

## Data Availability

All data produced are available online at GenBank (www.ncbi.nlm.nih.gov/genbank/) and GISAID (gisaid.org). Accession numbers are provided in the manuscript.

https://doi.org/10.5281/zenodo.8323328

## Acknowledgements

We gratefully acknowledge all authors from reference laboratories who submitted genome sequences to public databases. This study was partially supported by AMED under the grant number JP223fa627005.

## Contributions

**Gabriel Gonzalez**: data analysis and draft manuscript preparation; **Michael Carr**: genome sequencing and manuscript edition; **Tomás M. Kelleher**: genome sequencing; **Emer O’Byrne**: sample extraction, genome sequencing and manuscript edition; **Weronika Banka**: sample extraction, genome sequencing and manuscript edition; **Brian Keogan**: sample location and extraction, manuscript edition; **Charlene Bennett**: protocols validation; **Geraldine Franzoni**: protocols validation; **Patrice Keane**: genome sequencing and manuscript edition; **Luke W. Meredith**: genome sequencing protocol; **Nicola Fletcher**: genome sequencing and manuscript edition; **Jose Maria Urtasun-Elizari**: sequence analysis and manuscript preparation; **Jonathan Dean**: data collection and analysis, manuscript edition; **Brendan Crowley**: sample collection, results validation and manuscript edition; **Derval Igoe**: data evaluation and manuscript edition; **Eve Robinson**: data evaluation and manuscript edition; **Greg Martin**: data evaluation and manuscript edition; **Jeff Connell**: protocols validation; **Daniel Hare**: study oversight, manuscript preparation; **Cillian F. De Gascun**: study oversight, manuscript preparation. All authors reviewed the results and approved the final version of the manuscript.

