## Supplementary Materials for "Multiple Introductions of Mpox virus to Ireland during the 2022 International Outbreak"

This supplementary material is hosted as supporting information alongside the article [**Multiple Introductions of Mpox to Ireland during the 2022 International Outbreak**] by Gabriel Gonzalez *et al.*, on behalf of the authors, who remain responsible for the accuracy and appropriateness of the content. The same standards for ethics, copyright, attributions and permissions as for the article apply.

**Supplementary Table S1. Reference sequences from GISAID and GenBank considered for the analyses.**

Hosted in Zenodo.org as https://doi.org/10.5281/zenodo.8323328 (*Supplementary Table 1.xlsx*)

**Supplementary Table S2. Samples sequenced in the Republic of Ireland during the 2022 MPXV epidemic.**

Hosted in Zenodo.org as https://doi.org/10.5281/zenodo.8323328 (*Supplementary Table 2.xlsx*)

**Supplementary Table S3. Count of positions with mutations across the MPXV whole-genome sequences[+]**

| | | **TO** | | | |
| --- | --- | --- | --- | --- | --- |
| | | **A** | **T** | **G** | **C** |
| **FROM** | **A** | - | 1 | 3 | 2 |
| | **T** | 2 | - | 0 | 6 |
| | **G** | **132** | 3 | - | 2 |
| | **C** | 2 | **133** | 0 | - |

[+] These counts show the summary of positions with mutations to avoid double-counting any position due to the phylogenetic relations among the sequences.

**Supplementary Table S4. Distribution of mutations per coding regions.**

Hosted in Zenodo.org as https://doi.org/10.5281/zenodo.8323328 (*Supplementary Table 4.xlsx*)

* All non-synonymous mutations corresponded to amino acid substitutions

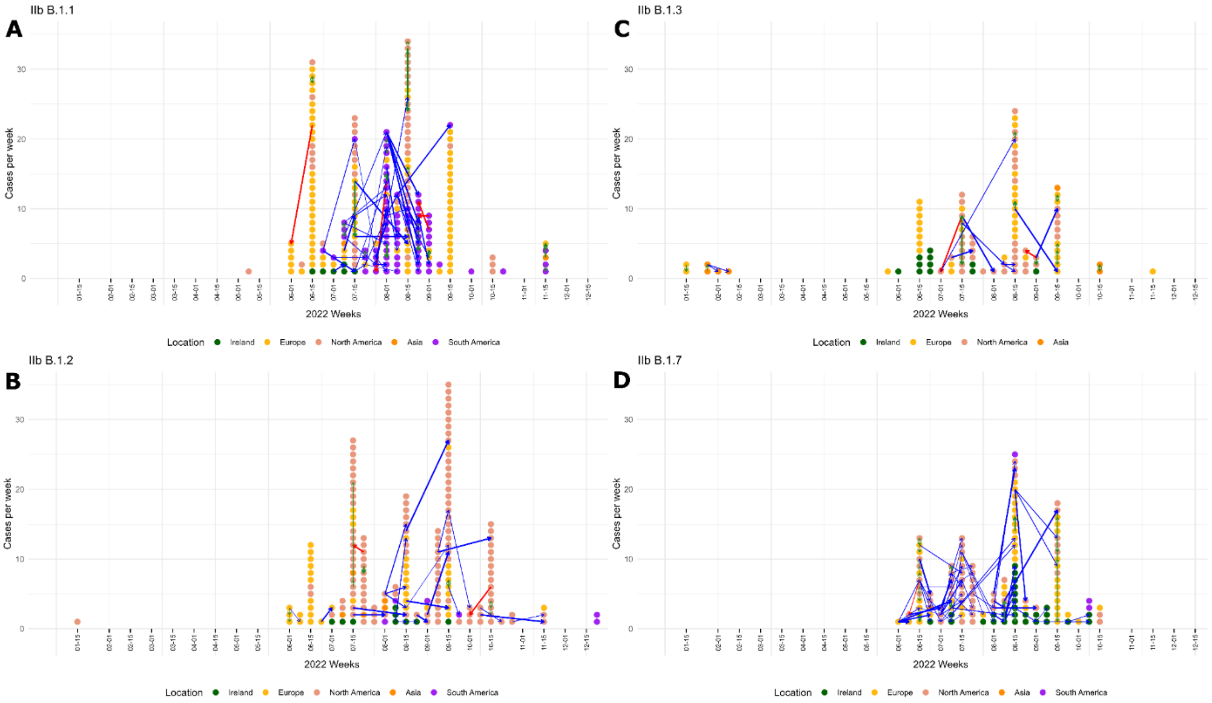


**Supplementary Figure S1. Inferred transmission between sequenced cases from different locations for the different MPXV subclades detected in the Republic of Ireland.** The clades with most cases per week (vertical axes) and the better supported inferred transmission (Outbreaker2 support >0.3 and two or less generations involved in the transmission) are shown along the weeks of 2022 (horizontal axes). Cases are coloured by location as summarised at the bottom of each panel. The transmissions are represented by arrows with width according to the support and coloured according to the direction as reverse, forward and in the same week, as red, blue and green, respectively. The presented clades are IIb **A)** B.1.1, **B)** B.1.2, **C)** B.1.3 and **D)** B.1.1.7.

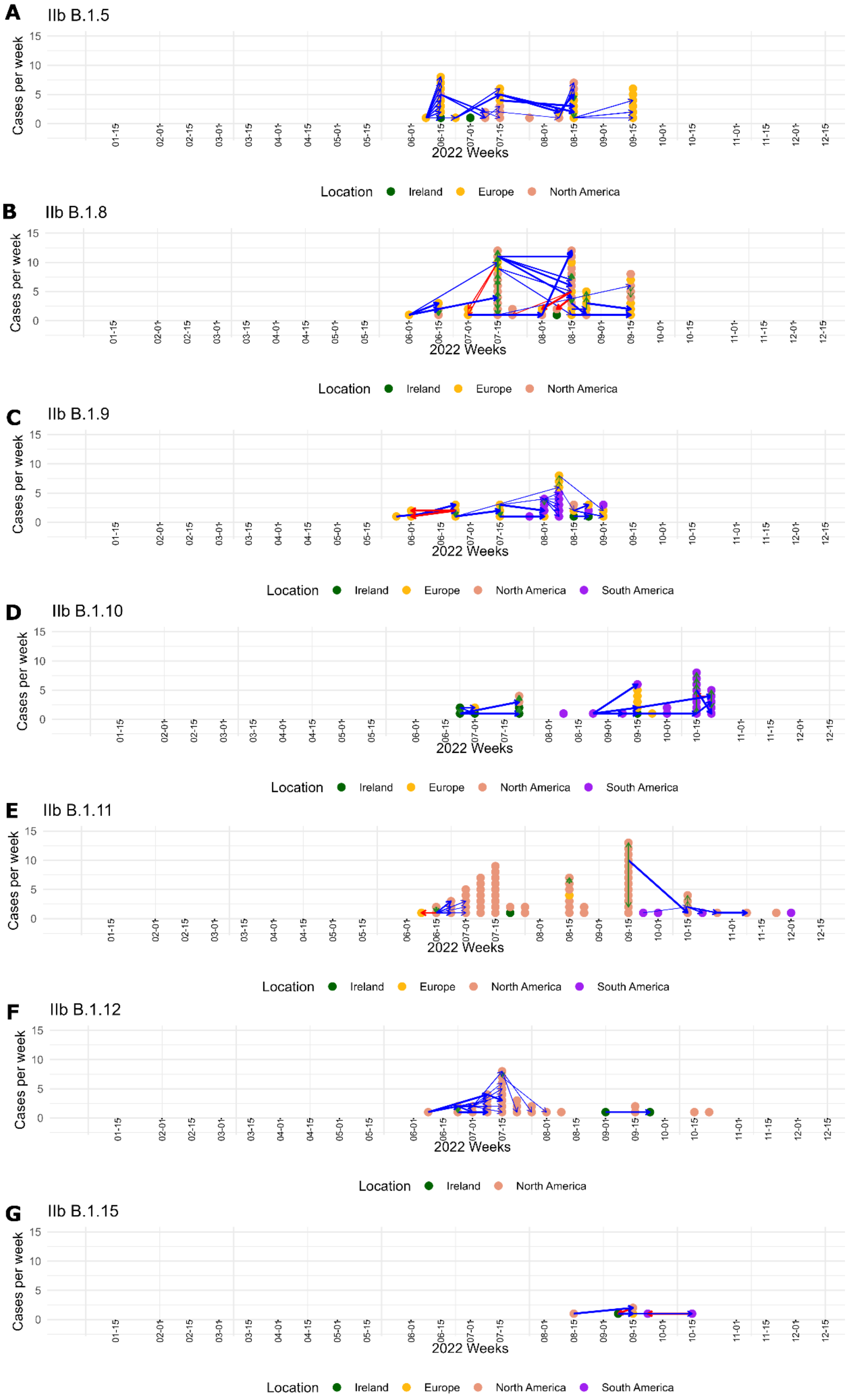


**Supplementary Figure S2. Inferred transmission between sequenced cases from different locations for the different subclades detected in the Republic of Ireland (Cont.).** (See Fig. 4) The clades with less cases were IIb **A)** B.1.5, **B)** B.1.8, **C)** B.1.9, **D)** B.1.10, **E)** B.1.11, **F)** B.1.12 and **G)** B.1.15.
